# Lipoprotein metabolism and inflammation in healthy young subjects – exploring the postprandial and postabsorptive phases following intake of a standardized meal

**DOI:** 10.1101/2025.05.15.25327690

**Authors:** Silje-Marie Jensen, Kirsten B. Holven, Stine M. Ulven, Åslaug Matre Anfinsen, Jutta Dierkes, Vegard Lysne, Jacob J. Christensen

**Author notes:** E-mails: Silje-Marie Jensen;, Kirsten B. Holven; Stine M. Ulven; Åslaug Matre Anfinsen; Jutta Dierkes; Vegard Lysne. Jacob J. Christensen (corresponding author).

## Abstract

**Background and aims:** Non-fasting blood sampling is increasingly recommended for routine lipid testing in cardiovascular disease risk assessment. However, these recommendations are mainly based on associations between lipid levels and self-reported time since the last meal in epidemiological studies. The aim of the present study was to explore the time-resolved variation in lipoproteins and inflammatory marker concentrations during the postprandial and postabsorptive period, in males and females ingesting a standard breakfast meal.

**Methods:** We used data from a trial where 34 normal-weighted subjects aged 20-30 y were included. Subjects fasted 12 hours, had blood sampled at baseline, then consumed a standardized breakfast meal, and had blood sampled 13 more times over the next 24 hours. NMR metabolomics was used to quantify lipoprotein subclasses and various biomarkers, and ELISA to analyze VCAM-1, ICAM-1, E-selectin, and IL-6. We characterized the postprandial and postabsorptive responses using visualizations and non-linear mixed effects models.

**Results:** Six VLDL subclasses increased by 11-429% within 2-4 hours postprandially before returning to baseline levels, followed by another increase 8-10 hours after intake of the breakfast meal. IDL and three LDL subclasses increased around 10-12% over 24 hours. Four HDL subclasses showed an inverse association with VLDL subclasses, reaching their lowest levels about 1 hour after the meal and peaking after 8-10 hours with a 5-15% increase from baseline. Participants had an average increase of 325% in IL-6 ten hours after breakfast, while other inflammatory biomarkers showed little change over time. For most biomarkers, males generally exhibited higher baseline concentrations compared to females, however the responses over time remained similar.

**Conclusions:** We observed small-to-moderate changes in plasma concentrations of most biomarkers in the postprandial and postabsorptive phases. Non-fasting lipid testing is likely a viable option for CVD risk assessment, as recommended by recent guidelines and consensus statements.

## Introduction

Blood concentrations of metabolites are not constant, but change dynamically throughout the day. For nutrition-related biomarkers, a major influencing factor is recent food intake, both in terms of food type, amount, and the time since ingestion of the meal. The time-point of blood sampling relative to food intake may be of importance when interpreting the results of a biochemical measure, as the concentrations of any biomarker may change considerably from food intake, and through the continuum that makes up the postprandial and postabsorptive (fasting) states [1].

Cardiovascular risk assessment includes the analysis and interpretation of a standard lipid panel [2]. Because triglycerides increase in the postprandial phase, lipids have traditionally been measured in fasting blood samples; however, non-fasting blood sampling is increasingly recommended for lipid testing [2–5]. Furthermore, inflammation is a key part of atherosclerosis, and biomarkers of low-grade inflammation have been suggested to improve risk classification compared to traditional risk factors alone [6–10]. Because inflammatory biomarkers may change during the postprandial and postabsorptive phases, the time-point of blood sampling may be important when interpreting the concentrations.

Non-fasting lipid testing may be preferred over fasting for several reasons [5]. For example, most people consume multiple meals and snacks throughout the day, meaning that the postprandial state dominates over a 24-hour period. Thus, non-fasting lipid profiles more accurately reflect the longer-term average concentration of plasma lipids and lipoproteins. Also, non-fasting blood samples are less burdensome for the patient and thus less susceptible to compliance issues, making them more cost-effective [5].

We recently investigated postprandial and postabsorptive changes for a standard lipid panel using clinical chemistry in a controlled setting with repeated samples [11]. We found that the concentrations of HDL-C and LDL-C decreased to approx. 2 hours (-4%) while TG peaked at 3 hours (+27%), which is in line with the epidemiological evidence. Recently, NMR metabolomics has been more often used to assess lipoprotein particles; however, NMR metabolomic-based lipoprotein particles and other metabolites are not yet characterized across the postprandial and postabsorptive phases.

Using state-of-the-art NMR metabolomics and ELISA analyses, we aimed to comprehensively describe the sex-specific dynamic variation in the concentrations of lipoproteins and inflammatory markers during the postprandial and postabsorptive phases. We used data from a recently conducted trial on 34 young and healthy subjects who had blood drawn after a 12 hour overnight fast, and then at 13 time points during the next 24 hours after ingesting a standardized breakfast meal.

## Subjects and methods

### Study design, setting and participants

In the present study, we investigated postprandial and postabsorptive changes in lipoprotein subclasses and inflammatory markers using data from a recently conducted trial. The study design has been described in detail previously [1].

Eligible participants were healthy men and women aged 20-30 years (birth years 1991-2001), with BMI 22-27 kg/m^2^, and without significant weight change (>5%) within three months before the study visit. Participants were recruited through social media and posters in the local area in Bergen, Norway.

In brief, participants consumed a standardized evening meal at 20:00 p.m. and then fasted overnight (approx. 12 hours). The following day, participants met at the Research Unit for Health Surveys at the University of Bergen, Norway, and had clinical measurements and blood samples taken at baseline. Participants were then instructed to consume a standardized breakfast in precisely 15 minutes. They were thereafter subjected to frequent blood sampling for 24 hours with a total of 13 additional blood samples drawn from each participant (12 blood samples first 12 hours while being supervised in the research unit, and one blood sample after non-supervised 12 hours at the following morning, after 24 hours). Fasting during the unsupervised hours was confirmed by increased ketone bodies measured throughout the study and after 24 hours [1].

During the 24 hours of blood sampling, the participants were fasting but free to drink water. The standardized meal was composed to mimic a habitual Norwegian breakfast and consisted of two slices of whole grain bread with butter and low-fat cheese, one slice of whole grain bread with strawberry jam, and one glass of orange juice. The composition of the meal was 520 kcal, of which 20,5 g (36 E%) was fat, 59,4 g (46 E%) was carbohydrates, 20,5 g (16 E%) was protein, and containing 5 g (1,9 E%) fiber.

### Assessment of lipoproteins with NMR metabolomics

Concentrations of lipoproteins were quantified by using high-throughput proton NMR spectroscopy metabolomics (2020 Algorithm; Nightingale Health, Helsinki, Finland). The platform provides 250 metabolic measures per sample of lipoprotein lipids, fatty acids, and small molecules such as amino acids, ketones, and glycolysis metabolites [12]. In the present work, we included 14 lipoprotein subclasses, the inflammatory marker Glycoprotein acetyls (GlycA), and lipids, cholesterol measurements, apolipoproteins, phospholipids, and lipid concentration within the subclasses. The lipoprotein subclasses were classified based on average particle sizes (in nanometers).

### Assessment of inflammatory response with ELISA

We used ELISA to analyze the inflammatory markers Vascular Cell Adhesion Molecule 1 (VCAM-1), Intercellular Adhesion Molecule 1 (ICAM-1), Endothelial Adhesion Molecule 1 (E-selection), and Interleukin 6 (IL-6). ELISA analyses were performed at the Department of Nutrition, UiO, using commercially available ELISA Duo and Quantikine kits (R&D Systems, Minneapolis, USA). The following kits were used: Human VCAM-1/CD106 DuoSet ELISA (DY809), Human ICAM-1/CD54 DuoSet ELISA (DY720), E-Selectin/CD62E DuoSet ELISA (DY724), Human IL-6 Quantikine HS ELISA (HS600C). Intra-assay CV was between 4.4-5.0 % for all assays.

### Statistical analyses

Data analyses were performed in R version 4.2.3 [13] using the RStudio IDE (https://posit.co/) and the tidyverse framework [14].

Data are presented with counts (and percentages) for categorical variables, or geometric mean (gMean) and geometric standard deviations (gSD) or geometric confidence intervals (gCI) for continuous variables. Results were presented as visualizations of metabolite concentrations as a function of time, with gMean and gCI superimposed on top of individual-level data. gCI was calculated by using the following formulas:

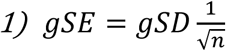

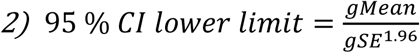

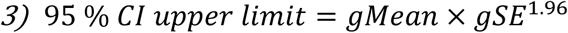

We also described the lowest and highest (peak) points on the curve trajectories, relative to the baseline level.

For each biomarker, we fitted non-linear mixed effect models using the lme4::lmer function with time, sex, BMI, and age as covariates, plus a random intercept for ID to account for dependency between each individual’s 14 data points. Time was modeled using a B-spline function using the splines::bs function, and the interaction terms for sex and time, age and time, and BMI and time were also included in the model. We extracted β coefficients, 95 % CIs and P values for the sex differences (males vs. females), and used analysis of variance (ANOVA) to derive global P values for the non-linear terms for the sex by time interaction (that is, the sex differences in response over time). Standard deviation (SD)-normalized coefficients and P values for sex are shown in the figures (normalized to enable comparison between panels), while full models on original scale are shown in Table S3, including coefficients and P values for BMI and age.

The sample size was calculated using an accuracy-in-parameter-estimation approach to estimate the majority of measurements withing a multiplicative margin-of-error of 10 %. The calculation are described in the main article [1].

### Ethical approvals

The present study was registered at ClinicalTrials.gov (NCT number 04989478), conducted in accordance with the Declaration of Helsinki, and approved by the Regional Committee for Research Ethics for the Western part of Norway, located at the University of Bergen (REK 236654). Participants signed a consent form prior to engaging in the study and had the chance to withdraw from the study at any time.

## Results

### Baseline characteristics

We included data from all 34 participants, of which 18 (52 %) were males and 16 (48%) females. 33 participants completed all measurements, and one participant (female) completed the first two hours (7 measurements). In total we received 469 blood samples. Compared to females, males were older (26.2 vs. 24.6 yrs) and had slightly higher BMI (23.9 vs. 23.1 kg/m^2^). Also, males had slightly higher levels of LDL cholesterol, while females had higher levels of HDL cholesterol (**Table 1**).

**Table 1.** Baseline characteristics of the study population.

|  | Overall,<br>N = 34 | Females,<br>N = 16<sup>a</sup> | Males,<br>N = 18 |
| --- | --- | --- | --- |
| **Clinical** |  |  |  |
| Age, yrs | 25.3 (1.1) | 24.5 (1.1) | 26.1 (1.1) |
| < 25.8 yrs | 17 (50%) | 9 (56%) | 8 (44%) |
| ≥ 25.8 yrs | 17 (50%) | 7 (44%) | 10 (56%) |
| Weight, kg | 73.4 (1.2) | 65.2 (1.1) | 80.9 (1.1) |
| Height, cm | 176 (1.1) | 168 (1.1) | 184 (1.0) |
| BMI, kg/m<sup>2</sup> | 23.5 (1.1) | 23.1 (1.1) | 23.8 (1.1) |
| < 25, kg/m<sup>2</sup> | 26 (79%) | 13 (87%) | 13 (72%) |
| ≥ 25, kg/m<sup>2</sup> | 7 (21%) | 2 (13%) | 5 (28%) |
| **Biochemical** |  |  |  |
| Glucose, mmol/l | 5.3 (1.1) | 5.1 (1.1) | 5.4 (1.1) |
| Insulin, mIU/L | 4.5 (1.9) | 4.4 (2.2) | 4.5 (1.8) |
| HbA1c, mmol/mol | 31 (1.1) | 31 (1.1) | 32 (1.1) |
| LDL-C, mmol/L | 2.5 (1.3) | 2.4 (1.2) | 2.6 (1.4) |
| HDL-C, mmol/L | 1.6 (1.2) | 1.7 (1.3) | 1.5 (1.2) |
| Triglycerides, mmol/L | 0.9 (1.5) | 0.8 (1.5) | 0.9 (1.5) |
| CRP, mg/L | 0.9 (2.6) | 1.4 (2.6) | 0.6 (2.1) |

Data are presented as geometric mean (geometric SD) for continuous variables and as count (percentage) for categorical variables. gSD indicates how much the data varies multiplicative from the gMean; for example, for overall age, most data are covered by the interval $[25.3/1.13 - 25.3 \times 1.13] = [22.4 - 28.6]$ yrs.
<sup>a</sup>n=15 for weight and BMI.
Abbreviations: BMI; body mass index, CRP; C-reactive protein, HbA1c; glycated hemoglobin, HDL-C; cholesterol in high-density lipoprotein, LDL-C; cholesterol in low-density lipoprotein.

### Standard lipid panel

We previously explored the postprandial and postabsorptive changes for a standard lipid panel using clinical chemistry; here, we present standard lipid panel measures analyzed by NMR metabolomics. Total-TG had the most pronounced changes during the 24-hour period (+18% at 3 hours, -21% at 10 hours), and HDL-C and ApoA1 showed a pattern that was the inverse of Total-TG (-5% at 1.5 hours, +9% at 12 hours) (**Figure 1**, **Tables S1** and **S2**). In contrast, LDL-C and correlated biomarkers showed minor changes in the early postprandial phase (first six hours) but increased slightly with increased time since the meal (+9% at 24 hours) (Figure 1). Responses were generally similar between sexes, although males seemed to show a larger response in the early postprandial phase (Figure 1).

**Figure 1.**
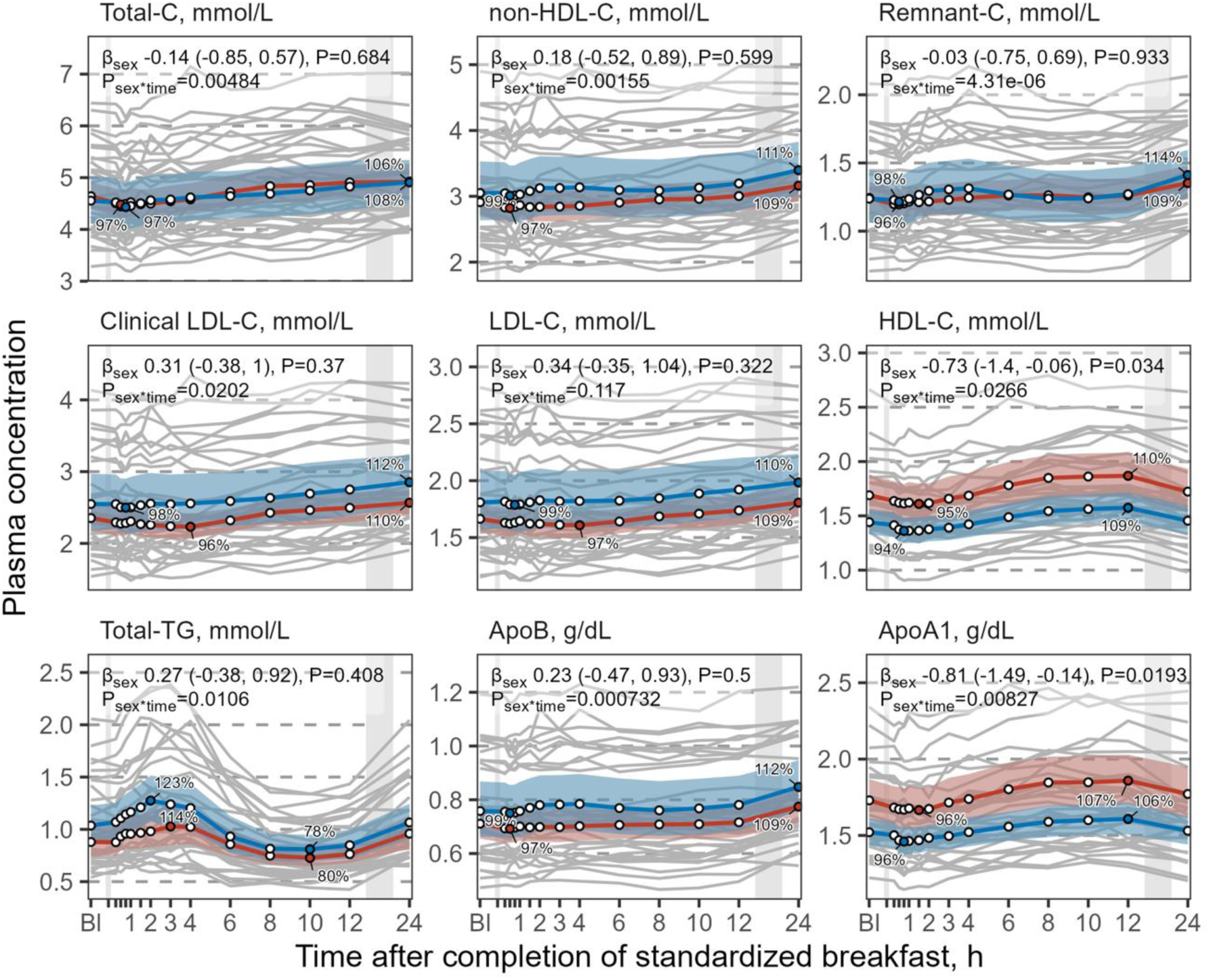
The plasma concentrations of standard lipid biomarkers as a function of time since completion of the standardized breakfast. The solid red and blue lines represent the geometric mean for females (n = 16) and males (n = 18), respectively, while the shaded areas represent the 95 % geometric confidence intervals (gCI). Individual levels are depicted as grey lines. The leftmost grey vertical line indicates the time of the standardized breakfast meal, while the rightmost grey vertical line indicates time spent outside the study center. Percentage of baseline are shown as average sex-specific minimum and maximum values. Sex differences (males compared to females) are shown as standard deviation (SD)-normalized β coefficients, 95 % CIs, and P values. Due to the SD-normalization, the coefficients can be compared across panels. The P values for the sex*time interaction are derived from an ANOVA of the underlying non-linear model (see Methods for model description). Abbreviations: apoB, apolipoprotein B; apoA1, apolipoprotein A1; Bl, baseline; C, cholesterol; h, hours; HDL, high density lipoprotein; LDL, low density lipoprotein; TG, triglycerides.

### Lipoprotein subclass particles and lipid content

The lipoprotein subclasses showed different plasma levels (**Figure 2**, Tables S1 and S2); for example, the gMean baseline concentrations were 0.0067 μM for L-VLDL-P, 0.64 μM for L-LDL-P (95-fold higher than L-VLDL-P), and 1.5 μM for L-HDL-P (223-fold higher than L-VLDL-P). Compared to females, males generally had higher levels of most VLDL and LDL particles, and lower levels of most HDL particles.

**Figure 2.**
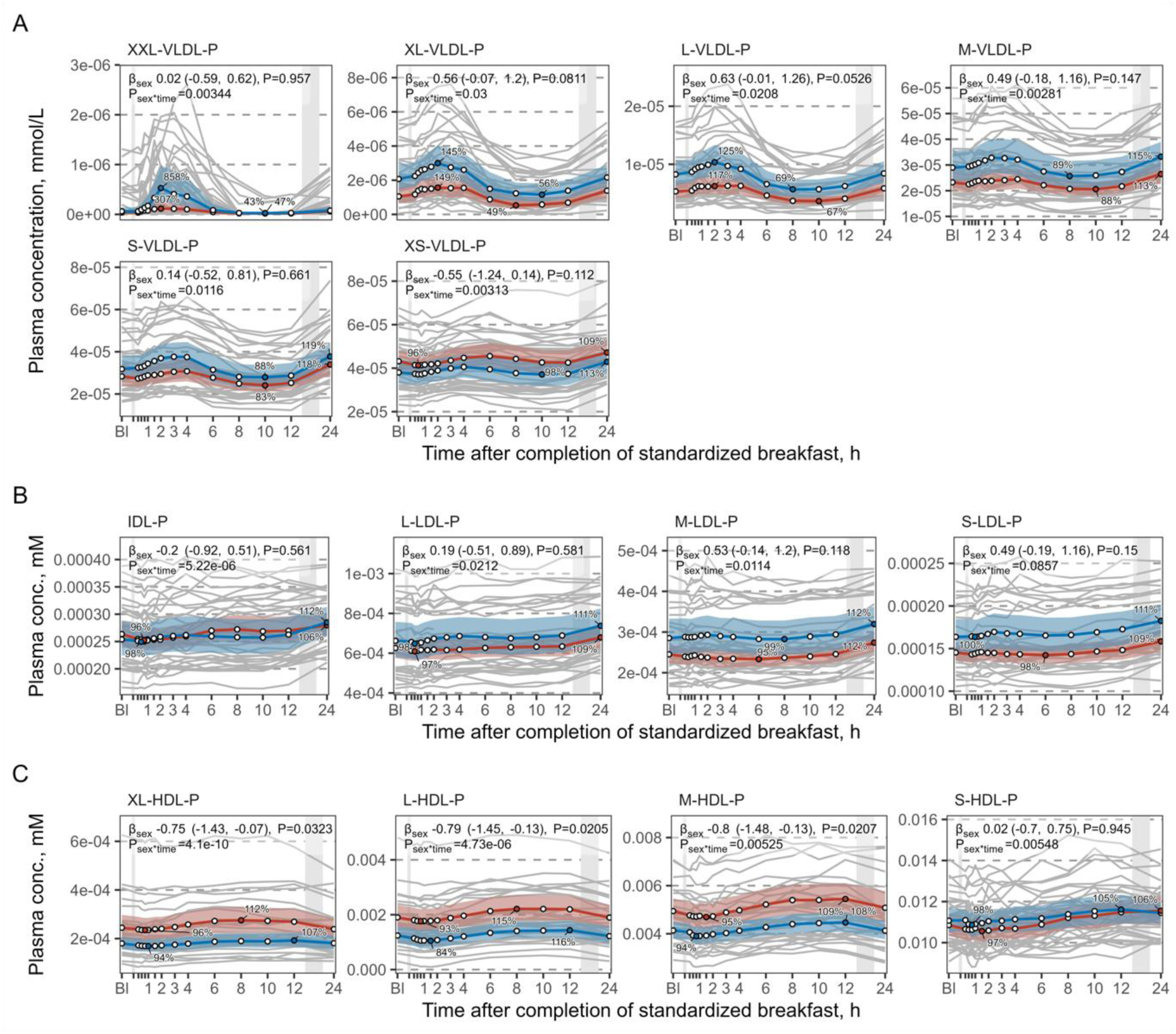
The plasma concentrations of lipoprotein subclass particles as a function of time since completion of the standardized breakfast. The figure shows VLDL particles (A), IDL and LDL particles (B) and HDL particles (C). See legend to Figure 1 for a comprehensive description. Abbreviations: Bl, baseline; HDL, high density lipoprotein; h, hours; IDL, intermediate density lipoprotein; L, large; LDL, low density lipoprotein; M, medium; mM, mmol/L; P, particle; S, small; VLDL, very low-density lipoprotein; XL, extra-large; XS, extra-small; XXL, extremely large.

The VLDL subclass particles (Figure 2A) and lipid content (Figure S1A) showed patterns similar to Total-TG (Figure 1). L-VLDL-P, for example, peaked at 2 hours (+21%) and were lowest at 8-10 hours (-32%), before increasing again towards 24 hours (+5%). While males had a slightly stronger higher VLDL increase in the early postprandial phase than females, responses were in general similar.

The IDL- and LDL subclass particles (Figure 2B) and lipid content (Figure S1B) were characterized by a pattern similar to Total- and LDL-C (Figure 1); for example, L-LDL-P showed minimal change in the postprandial phase but increased slightly in the fasting state (+10% at 24 hours) (Figure 2B). Females had higher LDL-TG and lower VLDL-TG. Responses were generally similar between sexes, except for IDL-P where sex-differences were observed despite absolute levels being numerically very similar.

For the HDL subclass particles (Figure 2C) and lipid content (Figure S1C), we observed a trajectory similar to HDL-C and ApoA1 (Figure 1). HDL-P, for example, decreased to the lowest concentration after 1 hour (-11%), and then peaked at 8-12 hours (+15%) before returning to baseline levels again. Interestingly, HDL-TG and LDL-TG had a trajectory resembling Total-TG and VLDL subclass particles. Responses were again fairly similar for males and females, but HDL-TG was higher in females compared to males.

### Inflammatory markers

We observed minimal changes for most of the inflammatory biomarkers (**Figure 3**). gMean (95 % CI) of baseline concentrations were 170 (150, 180) pg/mL for VCAM-1, 440 (410, 460) pg/mL for ICAM-1, 820 (740, 910) pg/mL for E-selectin, 1.2 (0.99, 1.5) pg/mL for IL-6, and 0.76 (0.73, 0.78) mmol/L for GlycA. Concentrations of GlycA, VCAM-1, and ICAM-1 exhibited a slight increase after 24 hours (+4-7%), while IL-6 exhibited the largest postprandial increase, with an average peak at 10 hours (+325%), before returning to baseline at 12-24 hours. There were no pronounced sex differences in response over time for the inflammatory biomarkers. GlycA was on average 0.8 SD lower among males compared to females.

**Figure 3.**
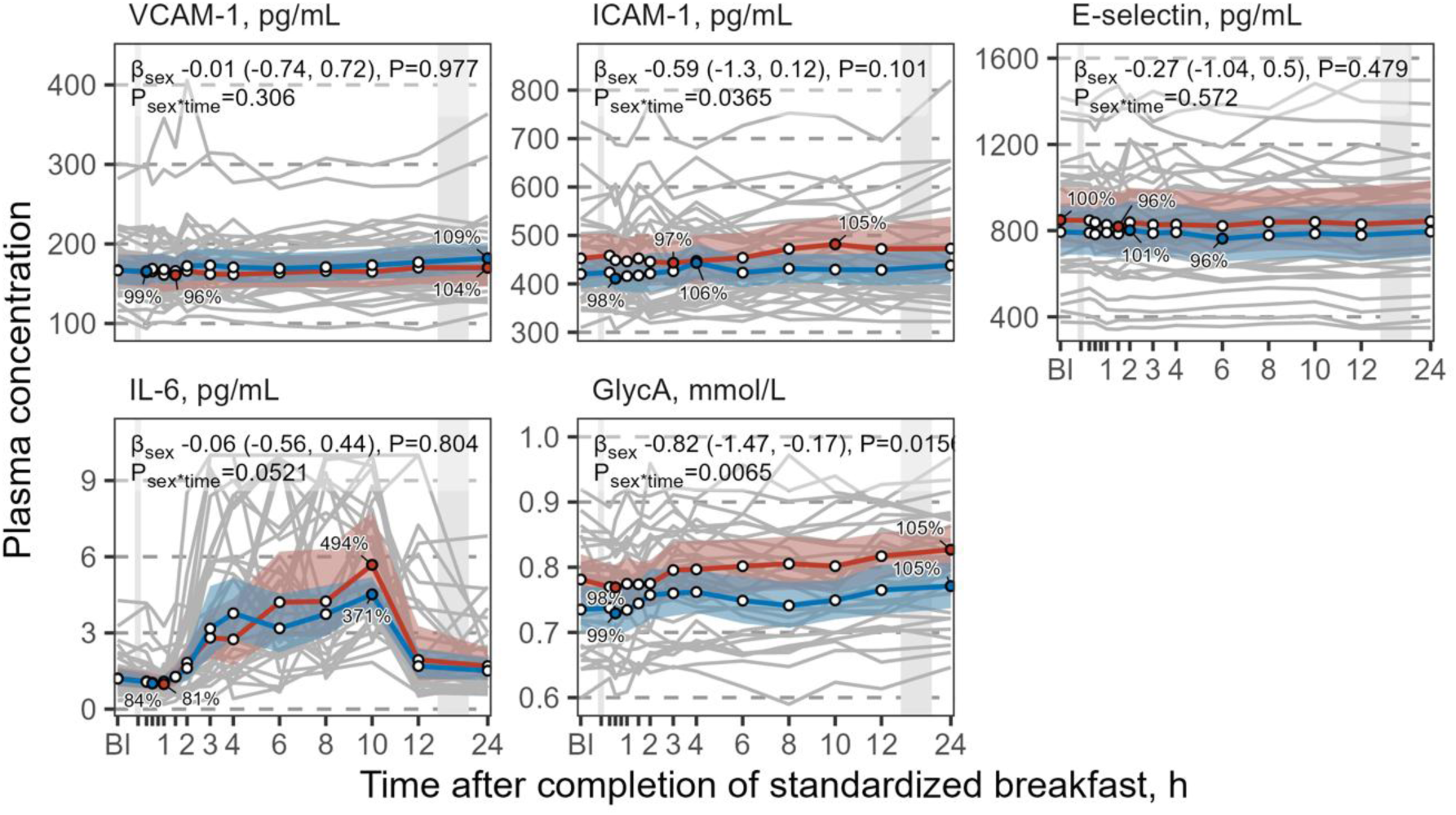
The plasma concentrations of inflammatory markers as a function of time since completion of the standardized breakfast. See legend to Figure 1 for a comprehensive description. ELISA data for timepoint number 4 (45 minutes after baseline) was skewed due to being run on a separate plate and were therefore removed from the data set. Abbreviation: Bl, baseline.

### Ketone bodies

Increased levels of ketone bodies confirmed that the participants were compliant to the study protocol. Ketone concentrations dropped rapidly following food intake (lowest point of -71% at 1.5h for bOHbutyrate for females and males combined, see Table S1 and S2), and subsequently increased substantially (peak of +1167% at 24h for bOHbutyrate) (**Figure S2**, Tables S1 and S2).

## Discussion

In the present study of healthy young adults, we conducted a comprehensive assessment of the postprandial and fasting lipoprotein metabolism and inflammatory response over 24 hours following intake of a standardized breakfast meal. Overall, food intake and fasting induced small-to-moderate changes in plasma concentrations of most biomarkers. There was prominent inter-individual variation in plasma concentrations; although responses were generally similar, there was some evidence of sex differences in response for some biomarkers. Our results support that non-fasting lipid testing is a viable option for CVD risk assessment, as recommended by recent guidelines and consensus statements.

Except for VLDL particles and TG, most markers remained stable throughout the 24-hour measuring period. Therefore, the timing of blood sampling may be more crucial for VLDL and TG than for the other markers. Similar changes in lipoproteins in the postprandial and postabsorptive phases are also reported by Bermingham et al. [15] and Hansson et al. [16], despite the subjects consuming a high-carbohydrate and/or high-fat breakfast.

Baseline plasma concentrations varied substantially between individuals; however, subjects tended to have similar response over time. Males and females generally had similar postprandial responses although we observed some small differences. Males generally had higher concentrations and larger responses than females; except in HDL particles, where females tended to have higher concentrations and larger responses. These observations were also observed by Bermingham *et al.* [15].

Several large-scale, population-based studies have shown that plasma lipids and lipoproteins (with the exception of triglycerides due to changes in VLDL particles) exhibit only modest postprandial changes and are considered clinically insignificant [4,5]. When comparing non-fasting and fasting lipid profiles, minor increases are typically found for plasma triglycerides, and slight decreases in total and LDL cholesterol levels, while HDL cholesterol levels are similar [4,5]. In the present study, we found that the observed absolute differences in concentrations across time-points were small, especially for biomarkers used in CVD risk assessment, such as TC, LDL-C, and apoB. The absolute changes were larger in TG and TG-correlated biomarkers; however, these are not used directly in CVD risk assessment.

Bermingham et al. also support that fasting status has a minor influence on plasma lipids [15]. Plasma lipid analyses assume that people adhere to normal dietary patterns with a variety of foods and nutrients, and normal meal patterns with several meals per day.

However, long-term fasting and fasting-mimicking diets are increasingly popular, and such diets may increase plasma lipid levels [3,17,18] and thus affect the risk estimation for cardiovascular diseases. Notably, we observed that several of the markers used to assess cardiovascular disease risk were highest after 24 hours of fasting, such as LDL-C and ApoB.

Taken together, our findings support that for comprehensive NMR metabolomics analyses of plasma lipids and lipoproteins, non-fasting or random blood sampling probably provides sufficient information for CVD risk assessment and thus most clinical decision-making.

Ingestion of food induces a physiological inflammatory response in the postprandial phase, likely driven in part by exposure to dietary lipids [9,19,20]. In our study, VCAM-1, ICAM-1, and E-selectin were unaffected in the postprandial and fasting period, which could be related to the size and the composition of the breakfast meal. Among previous studies on the postprandial effect of high-fat meals, some have reported increased levels of ICAM-1 [21,22], while others report no change over time for either of these biomarkers [23,24].

GlycA, a biomarker that reflect the overall level and complexity of the acute phase proteins, showed an unremarkable and modest increase with fasting [25], in line with previous findings by Mazidi *et al.* [20]. Taken together, for VCAM-1, ICAM-1, E-selectin and GlycA, we found no strong evidence that fasting status affects their plasma level to any meaningful degree [11].

In contrast and interestingly, IL-6 showed a remarkably higher level (+325% at 10 hours) after the meal compared to the fasting state at baseline. IL-6 has previously been reported to increase with age and following food intake, especially high-fat meals [19]. Also, exercise induces an increase in IL-6, especially endurance exercise [26,27]. Plasma IL-6 levels are important as they associate with future cardiovascular events in the general population both in observational studies [7,28,29] and mendelian randomization studies [30]; also, IL-6 receptor antagonism by tocilizumab increased myocardial salvage in patients with acute ST Elevation Myocardial Infarction [31], potentially driven by inhibition of the downstream mediator CXCL10 [32]. The evidence for IL-6 being causally involved in atherosclerosis is thus accumulating. It is possible that the changes observed in the present study in the postprandial and postabsorptive phase would have reclassified subjects with respect to risk [33].

Taken together, the inflammatory response to diet is variable across biomarkers, but the relevance to CVD risk assessment and clinical decision-making is still unclear. Time of blood sampling may be of importance for certain inflammatory biomarkers, such as IL-6, although this must be further studied.

### Strengths and limitations

The main strengths of the current study are the repeated sampling within individuals, including both males and females, collected in a well-controlled environment. The relatively homogenous study population, and standardized procedures, limits the external sources of variation such as due to age, health status, body composition, or physical activity. Further, the high frequency of samples provides accurate data on the dynamic changes in metabolite concentrations during the study period, and the sample size was large enough to achieve sufficient precision. A comprehensive panel of lipid and inflammatory biomarkers allow for a detailed description of the dynamics of the postprandial and postabsorptive lipid metabolism and inflammation. The compliance was good, as evidenced by the expected patterns in ketone, insulin, and glucose concentrations [1]. Furthermore, a strict time control limited any effect of day variation.

Some limitations warrant attention. Despite measures taken to limit variations due to other factors and maximize the internal validity, there were still some variations regarding body composition and estimated resting metabolic rate. These may have affected the postprandial responses and introduced some variability. Further, the standardized design and the artificial setting limits the external validity, and our ability to generalize to other contexts such as other age groups, non-healthy individuals, other meal composition or portion sizes, different prior food intake [34], and meals at other times of the day (circadian rhythm). For example, It is well known that diabetes may lead to disturbed lipid metabolism characterized by delayed concentration peak and prolonged high concentration of for example plasma triglycerides and VLDL particles [35]. However, our results may be applicable to the general healthy population across life-stage groups, supported by prior evidence demonstrating similar relative changes over time [3,36].

Although we have many measurements, especially in the early postprandial phase, we lack measurement points between 12 and 24 hours, which limits our resolution in this part of the fasting period. Also, hormonal changes in females due to menstrual cycle were not considered.

### Conclusions

In conclusion, whether food intake prior to blood sampling is of importance depends entirely on which biomarkers you aim to measure. As most lipids, except TG and VLDL, are relatively stable during the 24h following food intake, our data support the recommendation of non-fasting and random blood sampling as a viable option for CVD risk assessment (**Figure 4**). For inflammatory markers, we observed a substantial increase in IL-6 during the first 10h after food intake. This suggests that inflammatory markers should be measured after an overnight fast to limit the impact of prior food intake.

**Figure 4.**
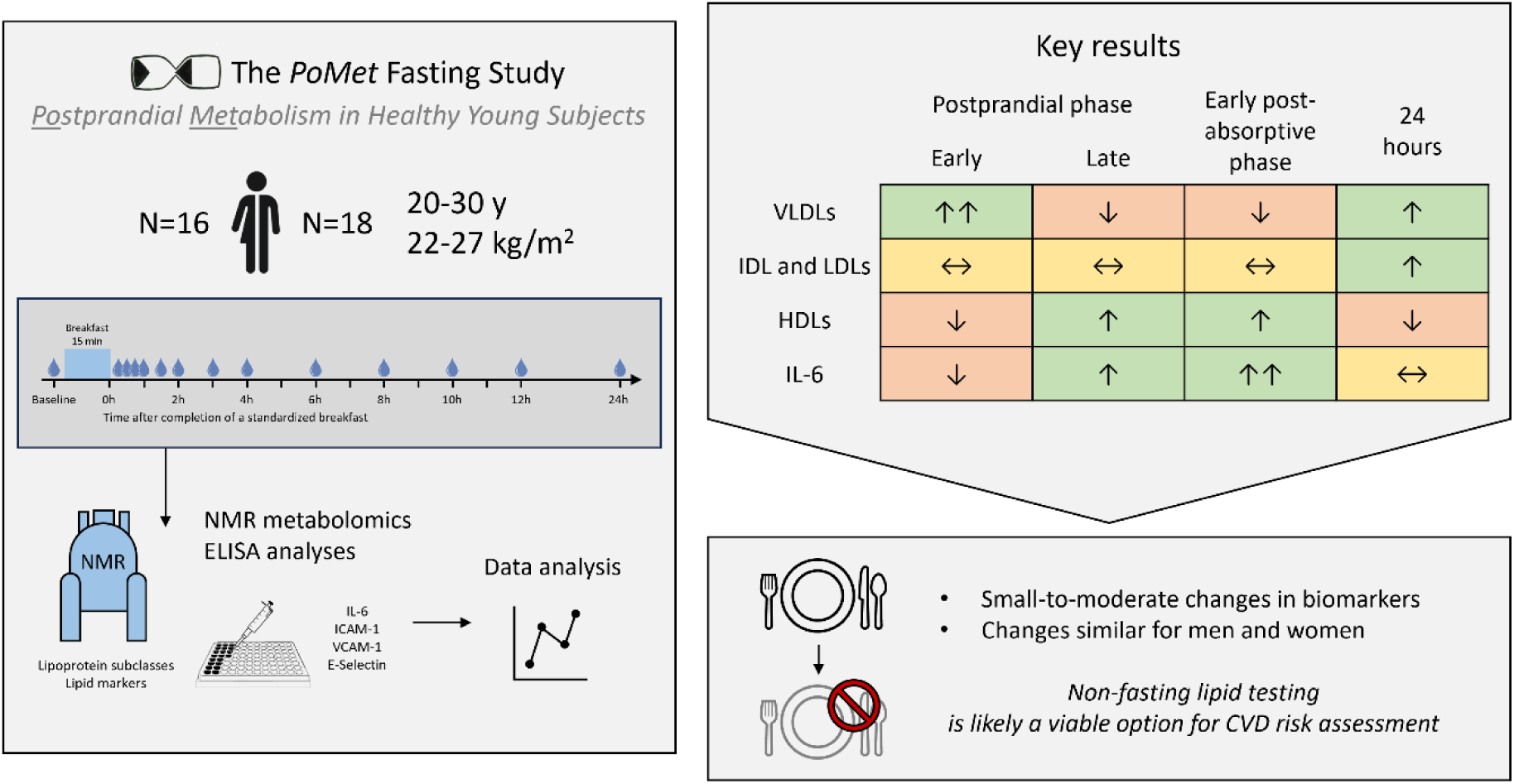
Graphical abstract. Time-resolved variation in lipoprotein subclasses and inflammatory biomarkers following intake of a standardized breakfast in healthy young adults. Abbreviations: CVD, cardiovascular disease; ELISA, enzyme-linked immunosorbent assay; h, hours; HDL, high-density lipoprotein; ICAM-1, intercellular adhesion molecule-1; IL-6, interleukin-6; IDL, intermediate-density lipoprotein; LDL, low-density lipoprotein; NMR, nuclear magnetic resonance; VCAM-1, vascular cell adhesion molecule-1; VLDL, very low-density lipoprotein; y, years.

## Data Availability

All data produced in the present work are contained in the manuscript.

## Abbreviations

BMI: body mass index
CE: cholesteryl esters
C: cholesterol
CVD: cardiovascular disease
ELISA: enzyme-linked immunosorbent assay
FC: free cholesterol
GlycA: glycoprotein acetyls
HDL: high-density lipoprotein
ICAM-1: intercellular adhesion molecule-1
IL-6: interleukin-6
LDL: low-density lipoprotein
L: large
M: medium
NMR: nuclear magnetic resonance
P: particles
PL: phospholipids
S: small
TG: triglycerides
VLDL: very-low-density lipoprotein
VCAM-1: vascular cell adhesion molecule-1
XXL: extremely large
XL: very large

## CRediT author statement

Silje-Marie Jensen: Conceptualization, Formal analysis, Investigation, Methodology, Validation, Visualization, Writing – original draft, Writing – review & editing.

Kirsten B. Holven: Conceptualization, Resources, Supervision, Validation, Writing – review & editing.

Stine M. Ulven: Conceptualization, Resources, Supervision, Validation, Writing – review & editing.

Åslaug Matre Anfinsen: Data curation, Investigation, Methodology, Project administration, Validation, Writing – review & editing.

Jutta Dierkes: Funding acquisition, Investigation, Methodology, Resources, Supervision, Validation, Writing – review & editing.

Vegard Lysne: Data curation, Funding acquisition, Investigation, Methodology, Project administration, Resources, Supervision, Validation, Writing – review & editing.

Jacob J. Christensen: Data curation, Formal analysis, Investigation, Methodology, Project administration, Resources, Software, Supervision, Validation, Visualization, Writing – original draft, Writing – review & editing.

## Acknowledgements

We thank all the study participants, and the fieldworkers at the Research Unit for Health Surveys.

## Declaration of generative AI and AI-assisted technologies in the writing process

During the preparation of this work the authors used GPT UiO in order to improve readability and language. After using this tool, the authors reviewed and edited the content as needed and take full responsibility for the content of the publication.

---

*Table S1. Absolute concentrations of all variables*

*Geometric mean and 95 % CI for absolute concentrations, for all variables, across all 14 measurements*. See the separate Excel document due to size.

*Table S2. Relative concentrations of all variables*

*Geometric mean and 95 % CI for level relative to baseline (scaled to 100), for all variables, across all 14 measurements*. See the separate Excel document due to size.

*Table S3. Model results for all variables*

*Model results for all variables including beta coefficients and 95 % CI for average differences, and ANOVA P values for interactions with spline term for time*. See the separate Excel document due to size.

**Figure S1.**
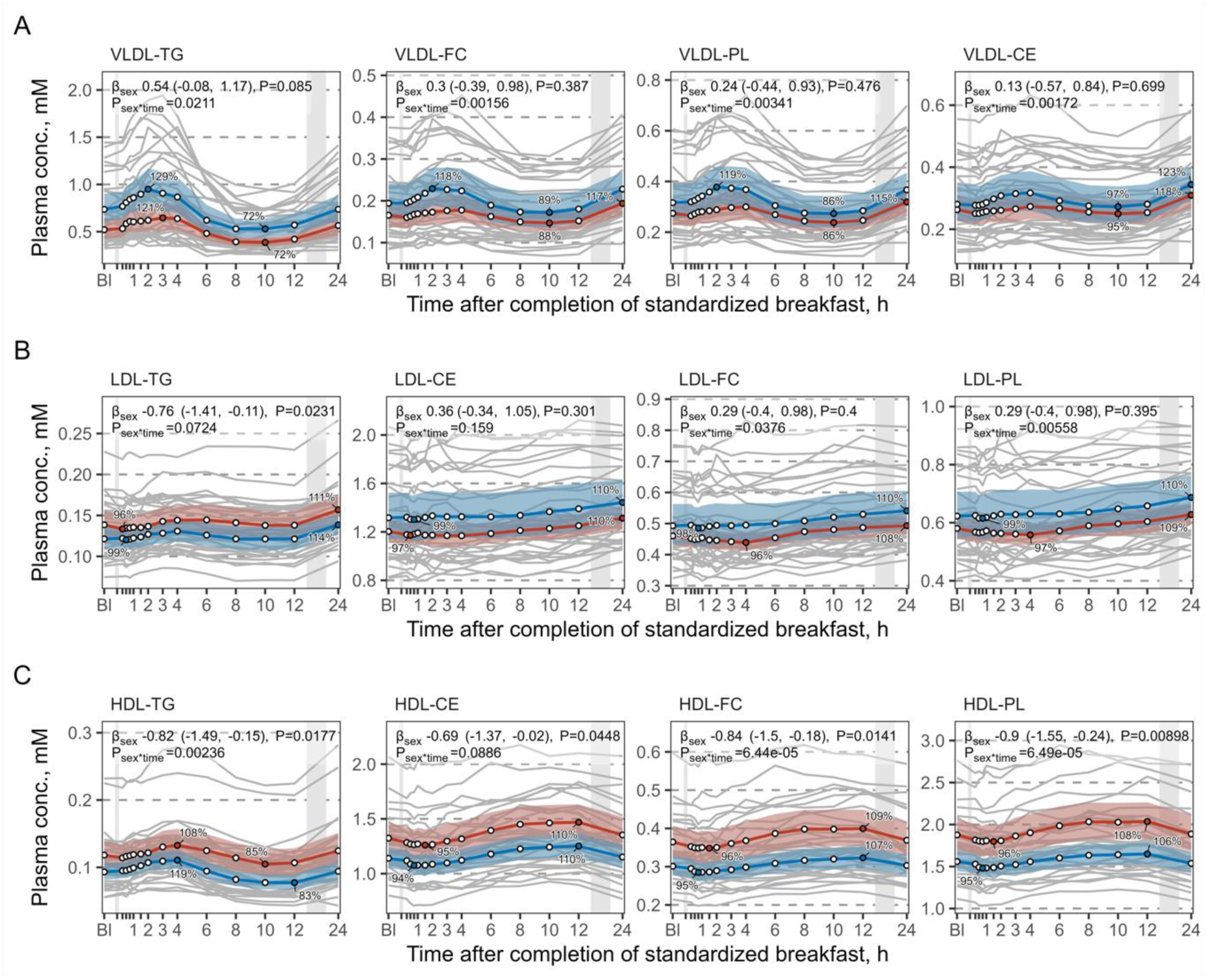
The plasma concentrations of main lipid types as a function of time since completion of the standardized breakfast. The solid red and blue lines represent the geometric mean for females (n = 16) and males (n = 18), respectively, while the shaded areas represent the 95 % geometric confidence intervals (CI). Individual levels are depicted as grey lines. The leftmost grey vertical line indicates the time of the standardized breakfast meal, while the rightmost grey vertical line indicates time spent outside the study center. Percentage of baseline are shown as average sex-specific minimum and maximum values. Sex differences (males compared to females) are shown as standard deviation (SD)-normalized β coefficients, 95 % CIs, and P values. Due to the SD-normalization, the coefficients can be compared across panels. The P values for the sex*time interaction are derived from an ANOVA of the underlying non-linear model (see Methods for model description). Abbreviations: Bl, baseline; CE, cholestetylester; FC, free cholesterol; HDL, high density lipoprotein; IDL, intermediate density lipoprotein; L, large; LDL, low density lipoprotein; M, medium; mM, mmol/L; P, particle; PL, phospholipids; S, small; TG, triglycerides; VLDL, very low-density lipoprotein; XL, extra-large; XS, extra-small; XXL, extremely large.

**Figure S2.**
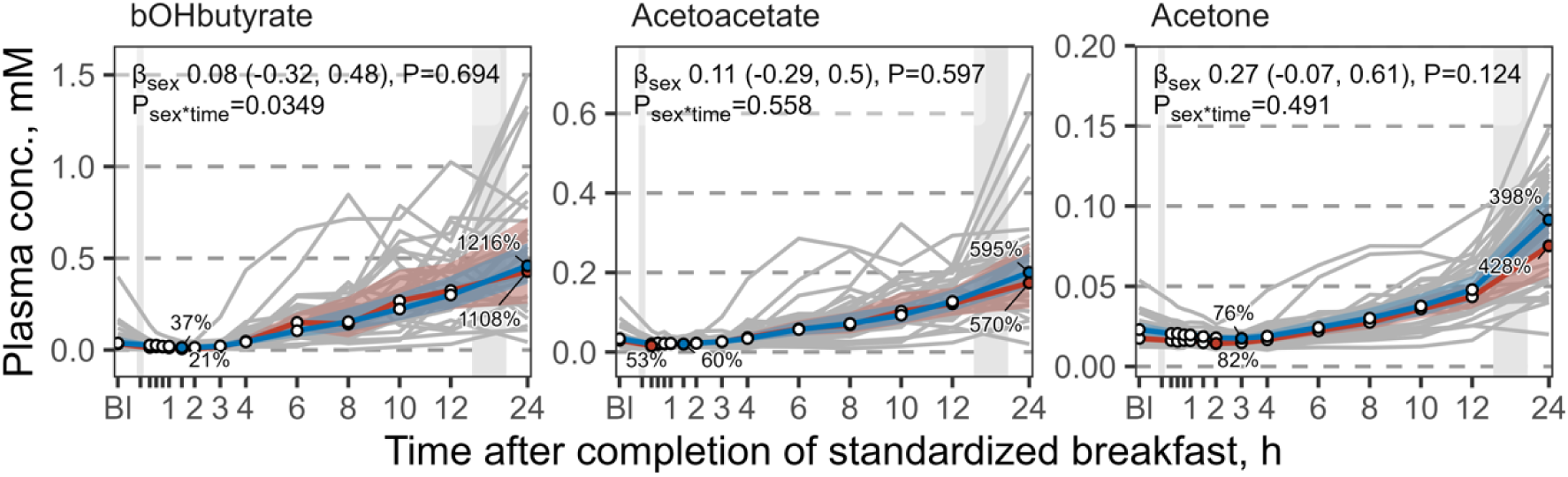
The plasma concentrations of ketone bodies as a function of time since completion of the standardized breakfast. The solid red and blue lines represent the geometric mean for females (n = 16) and males (n = 18), respectively, while the shaded areas represent the 95 % geometric confidence intervals (CI). Individual levels are depicted as grey lines. The leftmost grey vertical line indicates the time of the standardized breakfast meal, while the rightmost grey vertical line indicates time spent outside the study center. Percentage of baseline are shown as average sex-specific minimum and maximum values. Sex differences (males compared to females) are shown as standard deviation (SD)-normalized β coefficients, 95 % CIs, and P values. Due to the SD-normalization, the coefficients can be compared across panels. The P values for the sex*time interaction are derived from an ANOVA of the underlying non-linear model (see Methods for model description). Abbreviations: Bl, baseline; mM; millimole per liter.

## References

[1] Å.M. Anfinsen, C.O. Johannesen, V.H. Myklebust, H. Rosendahl-Riise, A. McCann, O.K. Nygård, J. Dierkes, V. Lysne, Time-resolved concentrations of serum amino acids, one-carbon metabolites and B-vitamin biomarkers during the postprandial and fasting state: the Postprandial Metabolism in Healthy Young Adults (PoMet) Study, Br. J. Nutr. (2023) 1–15. 10.1017/S0007114523002490.

[2] Cardiovascular disease: risk assessment and reduction, including lipid modification, National Institute for Health and Care Excellence (NICE), London, 2023. http://www.ncbi.nlm.nih.gov/books/NBK554923/ (accessed March 12, 2025).

[3] Keith N. Frayn, Rhys Evans, Human Metabolism: A Regulatory Perspective, 4th ed., Wiley-Blackwell, 2019.

[4] B.G. Nordestgaard, A. Langsted, S. Mora, G. Kolovou, H. Baum, E. Bruckert, G.F. Watts, G. Sypniewska, O. Wiklund, J. Borén, M.J. Chapman, C. Cobbaert, O.S. Descamps, A. von Eckardstein, P.R. Kamstrup, K. Pulkki, F. Kronenberg, A.T. Remaley, N. Rifai, E. Ros, M. Langlois, European Atherosclerosis Society (EAS) and the European Federation of Clinical Chemistry and Laboratory Medicine (EFLM) joint consensus initiative, Fasting is not routinely required for determination of a lipid profile: clinical and laboratory implications including flagging at desirable concentration cut-points-a joint consensus statement from the European Atherosclerosis Society and European Federation of Clinical Chemistry and Laboratory Medicine, Eur. Heart J. 37 (2016) 1944–1958. 10.1093/eurheartj/ehw152.

[5] A. Langsted, B.G. Nordestgaard, Worldwide Increasing Use of Nonfasting Rather Than Fasting Lipid Profiles, Clin. Chem. (2024) hvae046. 10.1093/clinchem/hvae046.

[6] P.M. Ridker, Clinical application of C-reactive protein for cardiovascular disease detection and prevention, Circulation 107 (2003) 363–369. 10.1161/01.cir.0000053730.47739.3c.

[7] P.M. Ridker, J.G. MacFadyen, R.J. Glynn, G. Bradwin, A.A. Hasan, N. Rifai, Comparison of interleukin-6, C-reactive protein, and low-density lipoprotein cholesterol as biomarkers of residual risk in contemporary practice: secondary analyses from the Cardiovascular Inflammation Reduction Trial, Eur. Heart J. 41 (2020) 2952–2961. 10.1093/eurheartj/ehaa160.

[8] T. Doi, A. Langsted, B.G. Nordestgaard, Dual elevated remnant cholesterol and C-reactive protein in myocardial infarction, atherosclerotic cardiovascular disease, and mortality, Atherosclerosis 379 (2023) 117141. 10.1016/j.atherosclerosis.2023.05.010.

[9] N. Seyedsadjadi, R. Grant, The Potential Benefit of Monitoring Oxidative Stress and Inflammation in the Prevention of Non-Communicable Diseases (NCDs), Antioxid. Basel Switz. 10 (2020) 15. 10.3390/antiox10010015.

[10] P.M. Ridker, M.V. Moorthy, N.R. Cook, N. Rifai, I.-M. Lee, J.E. Buring, Inflammation, Cholesterol, Lipoprotein(a), and 30-Year Cardiovascular Outcomes in Women, N. Engl. J. Med. 391 (2024) 2087–2097. 10.1056/NEJMoa2405182.

[11] Å.M. Anfinsen, V.H. Myklebust, C.O. Johannesen, J.J. Christensen, J. Laupsa-Borge, J. Dierkes, O. Nygård, A. McCann, H. Rosendahl-Riise, V. Lysne, Serum concentrations of lipids, ketones and acylcarnitines during the postprandial and fasting state: the Postprandial Metabolism (PoMet) study in healthy young adults, Br. J. Nutr. 132 (2024) 851–861. 10.1017/S0007114524001934.

[12] P. Soininen, A.J. Kangas, P. Würtz, T. Suna, M. Ala-Korpela, Quantitative Serum Nuclear Magnetic Resonance Metabolomics in Cardiovascular Epidemiology and Genetics, Circ. Cardiovasc. Genet. 8 (2015) 192–206. 10.1161/CIRCGENETICS.114.000216.

[13] R Core Team (2023). R: A language and environment for statistical computing. R Foundation for Statistical Computing, Vienna, Austria. URL: https://www.R-project.org/., (n.d.).

[14] Wickham H, Averick M, Bryan J, Chang W, McGowan LD, Françoi R, Grolemun G, Haye A, Henr L, Heste J, Kuh M, Pederse TL, Mille E, Bach SM, Müll K, Oo, J, Robins, D, Seid, DP, Spi, V, Takahas, K, Vaugh, D, Wil, C, W, K, Yutani, H (2019). Welcome to the tidyverse. Journal of Open Source Software, 4(43), 1686. doi:10.21105/joss.01686 https://doi.org/10.21105/joss.01686, (n.d.).

[15] K.M. Bermingham, M. Mazidi, P.W. Franks, T. Maher, A.M. Valdes, I. Linenberg, J. Wolf, G. Hadjigeorgiou, T.D. Spector, C. Menni, J.M. Ordovas, S.E. Berry, W.L. Hall, Characterisation of Fasting and Postprandial NMR Metabolites: Insights from the ZOE PREDICT 1 Study, Nutrients 15 (2023) 2638. 10.3390/nu15112638.

[16] P. Hansson, K.B. Holven, L.K.L. Øyri, H.K. Brekke, G.O. Gjevestad, M. Thoresen, S.M. Ulven, Sex differences in postprandial responses to different dairy products on lipoprotein subclasses: a randomised controlled cross-over trial, Br. J. Nutr. 122 (2019) 780–789. 10.1017/S0007114519001429.

[17] N.G. Norwitz, W.C. Cromwell, Oreo Cookie Treatment Lowers LDL Cholesterol More Than High-Intensity Statin therapy in a Lean Mass Hyper-Responder on a Ketogenic Diet: A Curious Crossover Experiment, Metabolites 14 (2024) 73. 10.3390/metabo14010073.

[18] V.B. Matzkin, C. Geissler, R. Coniglio, J. Selles, M. Bello, Cholesterol concentrations in patients with Anorexia Nervosa and in healthy controls, Int. J. Psychiatr. Nurs. Res. 11 (2006) 1283–1293.

[19] S.R. Emerson, S.P. Kurti, C.A. Harms, M.D. Haub, T. Melgarejo, C. Logan, S.K. Rosenkranz, Magnitude and Timing of the Postprandial Inflammatory Response to a High-Fat Meal in Healthy Adults: A Systematic Review, Adv. Nutr. Bethesda Md 8 (2017) 213–225. 10.3945/an.116.014431.

[20] M. Mazidi, A.M. Valdes, J.M. Ordovas, W.L. Hall, J.C. Pujol, J. Wolf, G. Hadjigeorgiou, N. Segata, N. Sattar, R. Koivula, T.D. Spector, P.W. Franks, S.E. Berry, Meal-induced inflammation: postprandial insights from the Personalised REsponses to DIetary Composition Trial (PREDICT) study in 1000 participants, Am. J. Clin. Nutr. 114 (2021) 1028–1038. 10.1093/ajcn/nqab132.

[21] A. Ceriello, L. Quagliaro, L. Piconi, R. Assaloni, R. Da Ros, A. Maier, K. Esposito, D. Giugliano, Effect of postprandial hypertriglyceridemia and hyperglycemia on circulating adhesion molecules and oxidative stress generation and the possible role of simvastatin treatment, Diabetes 53 (2004) 701–710. 10.2337/diabetes.53.3.701.

[22] S. Marchesi, G. Lupattelli, R. Lombardini, A.R. Roscini, D. Siepi, G. Vaudo, M. Pirro, H. Sinzinger, G. Schillaci, E. Mannarino, Effects of fenofibrate on endothelial function and cell adhesion molecules during post-prandial lipemia in hypertriglyceridemia, J. Clin. Pharm. Ther. 28 (2003) 419–424. 10.1046/j.0269-4727.2003.00512.x.

[23] D. Rubin, S. Claas, M. Pfeuffer, M. Nothnagel, U.R. Foelsch, J. Schrezenmeir, s-ICAM-1 and s-VCAM-1 in healthy men are strongly associated with traits of the metabolic syndrome, becoming evident in the postprandial response to a lipid-rich meal, Lipids Health Dis. 7 (2008) 32. 10.1186/1476-511X-7-32.

[24] W.L. Hall, A. Alkoblan, P.S. Gibson, M. D’Annibale, A. Coekaerts, M. Bauer, J.H. Bruce, B. Lecomte, A. Penhoat, F. Laugerette, M.-C. Michalski, L.J. Salt, P.J. Wilde, S.E. Berry, Postprandial lipid and vascular responses following consumption of a commercially-relevant interesterified palmitic acid-rich spread in comparison to functionally-equivalent non-interesterified spread and spreadable butter: a randomised controlled trial in healthy adults, Food Funct. 15 (n.d.) 2733–2750. 10.1039/d3fo05324e.

[25] R.A. Ballout, A.T. Remaley, GlycA: A New Biomarker for Systemic Inflammation and Cardiovascular Disease (CVD) Risk Assessment, J. Lab. Precis. Med. 5 (2020) 17. 10.21037/jlpm.2020.03.03.

[26] L. Wallberg, C. Mikael Mattsson, J.K. Enqvist, B. Ekblom, Plasma IL-6 concentration during ultra-endurance exercise, Eur. J. Appl. Physiol. 111 (2011) 1081–1088. 10.1007/s00421-010-1737-7.

[27] T.M. Kistner, B.K. Pedersen, D.E. Lieberman, Interleukin 6 as an energy allocator in muscle tissue, Nat. Metab. 4 (2022) 170–179. 10.1038/s42255-022-00538-4.

[28] P.M. Ridker, N. Rifai, M.J. Stampfer, C.H. Hennekens, Plasma concentration of interleukin-6 and the risk of future myocardial infarction among apparently healthy men, Circulation 101 (2000) 1767–1772. 10.1161/01.cir.101.15.1767.

[29] S. Kaptoge, S.R.K. Seshasai, P. Gao, D.F. Freitag, A.S. Butterworth, A. Borglykke, E. Di Angelantonio, V. Gudnason, A. Rumley, G.D.O. Lowe, T. Jørgensen, J. Danesh, Inflammatory cytokines and risk of coronary heart disease: new prospective study and updated meta-analysis, Eur. Heart J. 35 (2014) 578–589. 10.1093/eurheartj/eht367.

[30] G. Ou, H. Cai, K. Yao, Z. Qiu, Y. Yang, Y. Chen, X. Chen, Exploring the therapeutic potential of interleukin-6 receptor blockade in cardiovascular disease treatment through Mendelian randomization, Sci. Rep. 14 (2024) 21452. 10.1038/s41598-024-72195-4.

[31] K. Broch, A.K. Anstensrud, S. Woxholt, K. Sharma, I.M. Tøllefsen, B. Bendz, S. Aakhus, T. Ueland, B.H. Amundsen, J.K. Damås, E.S. Berg, E. Bjørkelund, C. Bendz, E. Hopp, O. Kleveland, K.H. Stensæth, A. Opdahl, N.-E. Kløw, I. Seljeflot, G.Ø. Andersen, R. Wiseth, P. Aukrust, L. Gullestad, Randomized Trial of Interleukin-6 Receptor Inhibition in Patients With Acute ST-Segment Elevation Myocardial Infarction, J. Am. Coll. Cardiol. 77 (2021) 1845– 1855. 10.1016/j.jacc.2021.02.049.

[32] S. Prapiadou, L. Živković, B. Thorand, M.J. George, S.W. van der Laan, R. Malik, C. Herder, W. Koenig, T. Ueland, O. Kleveland, P. Aukrust, L. Gullestad, J. Bernhagen, G. Pasterkamp, A. Peters, A.D. Hingorani, J. Rosand, M. Dichgans, C.D. Anderson, M.K. Georgakis, Proteogenomic Data Integration Reveals CXCL10 as a Potentially Downstream Causal Mediator for IL-6 Signaling on Atherosclerosis, Circulation 149 (2024) 669–683. 10.1161/CIRCULATIONAHA.123.064974.

[33] M.S. Khan, K.M. Talha, M.H. Maqsood, J.A. Rymer, B.A. Borlaug, K.F. Docherty, A. Pandey, F. Kahles, M. Cikes, C.S.P. Lam, A. Ducharme, A.A. Voors, A.F. Hernandez, A.M. Lincoff, M.C. Petrie, P.M. Ridker, M. Fudim, Interleukin-6 and Cardiovascular Events in Healthy Adults: MESA, JACC Adv. 3 (2024) 101063. 10.1016/j.jacadv.2024.101063.

[34] M.D. Robertson, M. Parkes, B.F. Warren, D.J.P. Ferguson, K.G. Jackson, D.P. Jewell, K.N. Frayn, Mobilisation of enterocyte fat stores by oral glucose in humans, Gut 52 (2003) 834–839. 10.1136/gut.52.6.834.

[35] H. Yanai, H. Adachi, M. Hakoshima, H. Katsuyama, Postprandial Hyperlipidemia: Its Pathophysiology, Diagnosis, Atherogenesis, and Treatments, Int. J. Mol. Sci. 24 (2023) 13942. 10.3390/ijms241813942.

[36] Y. Nabeno, Y. Fukuchi, Y. Matsutani, M. Naito, Influence of aging and menopause on postprandial lipoprotein responses in healthy adult women, J. Atheroscler. Thromb. 14 (2007) 142–150. 10.5551/jat.14.142.

